# Adapting a Data to Care Approach for HIV to Promote HCV Cure for Persons with HIV/HCV Co-infection

**DOI:** 10.1101/2023.10.26.23297615

**Authors:** Maximilian Wegener, Deborah Gosselin, Ralph Brooks, Suzanne Speers, Merceditas Villanueva

## Abstract

With Direct Acting Antivirals for Hepatitis C virus (HCV), cure is possible in >95% including those with HIV/HCV co-infection. Achieving strategic targets for cure requires addressing barriers including suboptimal care engagement. We adapted Data to Care (D2C), a public health strategy designed to identify and link persons out of care (OOC) for HIV, for persons with HIV/HCV co-infection untreated for HCV. In partnership with Connecticut Department of Public Health (DPH), persons OOC for HIV (defined as no HIV surveillance laboratory tests from 10/1/2018-10/1/2019) were matched to a list of persons co-infected with HIV/HCV (through 12/31/2019). We used a three-phase follow-up approach (pre-work, case conferencing, and Disease Intervention Specialist (DIS) follow-up) to track outreach outcomes and re-engagement/HCV cure success. There were 90 HIV/HCV co-infected persons who were OOC for HIV. The pre-work and case conferencing phases determined that 33 (36.7%) had previous HCV cure or were in treatment. There were 41 eligible for DIS-follow-up of which 21 (51%) were successfully contacted and 7 (33%) successfully re-engaged (kept appointment with HCV provider). No new HCV treatment initiations were recorded. Using a D2C approach, we identified and conducted outreach to persons who were OOC for HIV to promote HCV treatment. This approach resulted in intensive data clean-up and outreach efforts which produced modest re-engagement and no HCV treatment initiations. Future studies should develop alternative and complementary interventions to promote effective re-engagement and HCV treatment.

## INTRODUCTION

Historically, viral hepatitis, including Hepatitis C (HCV), has been the leading cause of chronic liver disease-related death globally.^1^ Persons co-infected with HIV have increased risk of liver-related complications compared to persons with mono-infection.^2–4^ The global burden of co-infection is estimated at 2.3 million (6.2% of prevalent HIV cases) with persons who inject drugs (PWID) having the greatest risk.^2–5^ HCV co-infection prevalence estimates in the U.S. vary by location and HIV risk factors.^6–10^ Direct acting antiviral (DAA) drugs with >95% cure rates for HCV have been extended to those with HIV/HCV co-infection, prompting strategic goal setting by the World Health Organization (WHO) to cure 80% of persons with diagnosed HCV infection by 2030.^5, 11–12^ The Centers for Disease Control and Prevention (CDC) has developed a strategic plan for the U.S. aimed at increasing HCV cure for chronically infected persons to ≥85% by 2030.^13^

Simplified treatment algorithms have been proposed to expand HCV treatment in uncomplicated patients (e.g., treatment naive, compensated cirrhosis).^14–15^ A global study looking at simplified implementation using a minimal monitoring approach showed sustained virologic response (SVR) rates of >95% including for persons HIV/HCV co-infected.^14–15^ The rollout of DAAs has resulted in population level success and in individual clinics, SVR rates ranging from 50% to 75% have been reported.^16–20^ Nonetheless, gaps in treatment persist due to system, clinic, and patient level issues. The optimal approach for identifying and engaging untreated persons into HCV treatment is not known.

Data to Care (D2C), a public health strategy originally designed using HIV surveillance and other data sources to identify and link newly diagnosed or to re-engage out-of-care (OOC) PWH, is one possible way to address this issue. The CDC’s HIV/AIDS Strategic Plan has advocated expanding D2C programs for PWH who have fallen OOC to improve the HIV continuum of care. A CDC-funded randomized control trial (RCT) found health-department employed disease intervention specialists (DIS) were more likely to re-engage newly OOC (i.e. 6 months without HIV laboratory tests or clinic visit) PWH at 90 days compared to the clinics’ standard of care.^21^

We hypothesized that OOC PWH co-infected with HCV were unlikely to have undergone DAA treatment due to lack of HIV care engagement and could benefit from this D2C approach. In this study, we partnered with the Connecticut Department of Public Health (CT DPH) to identify HIV OOC persons co-infected with HCV and piloted a re-engagement strategy including working with DIS, to improve the HCV viral clearance cascade.^22–23^

## METHODS

This study was part of a HRSA Special Project of National Significance (SPNS 047) initiative entitled “Curing Hepatitis C Among People of Color Living with HIV” which was awarded to Yale School of Medicine and conducted in partnership with the CT DPH. The project goal was to assess and promote efforts to treat HCV among HIV/HCV co-infected persons with a secondary goal of improving partnerships between the CT DPH and individual clinics.

### Surveillance Data Sources

Two CT DPH surveillance databases were used: the enhanced HIV/AIDS Reporting System (eHARS) for HIV and the CT Electronic Disease Surveillance System (CTEDSS) for HCV. The CDC developed eHARS for public health agencies to collect and report HIV surveillance data.^23–24^ The CT DPH uses CTEDSS as their notifiable disease repository for CDC-required reporting. In CT, HIV data in eHARS dates to 1981 and CTEDSS recorded data beginning in 1994. CTEDSS includes HCV antibody (positive only) and polymerase chain reaction (PCR) (positive and negative) results. Negative PCR results have been reported through electronic lab reporting (ELR) since 2016. Roughly 80% of all HCV laboratory results in CT are reported through ELR.

### Inclusion Criteria for Data to Care Intervention

The Data to Care (D2C) intervention focused on persons with HIV/HCV co-infection who were out of care (OOC) for HIV and had untreated HCV. **Figure 1** outlines the flow of surveillance data for determining eligibility for the intervention. First, the CT DPH epidemiologist created an HIV/HCV co-infected list by matching eHARS (from 1/1/2009 to 8/6/2019) and CTEDSS (from 1/1/1994 to 8/17/2019) using a CDC developed hierarchical deterministic matching program in SAS 9.4.^9^ Matching keys consisting of identifiable variables to determine persons present in both surveillance systems including: first name, last name, date of birth, and social security number.^25^

We chose the greater duration of time for CTEDSS to capture the largest possible pool of HCV data available. We chose the 10-year timeframe for eHARS to optimize the list of PWH who were more likely to be alive and living in CT.

**Figure 1:**
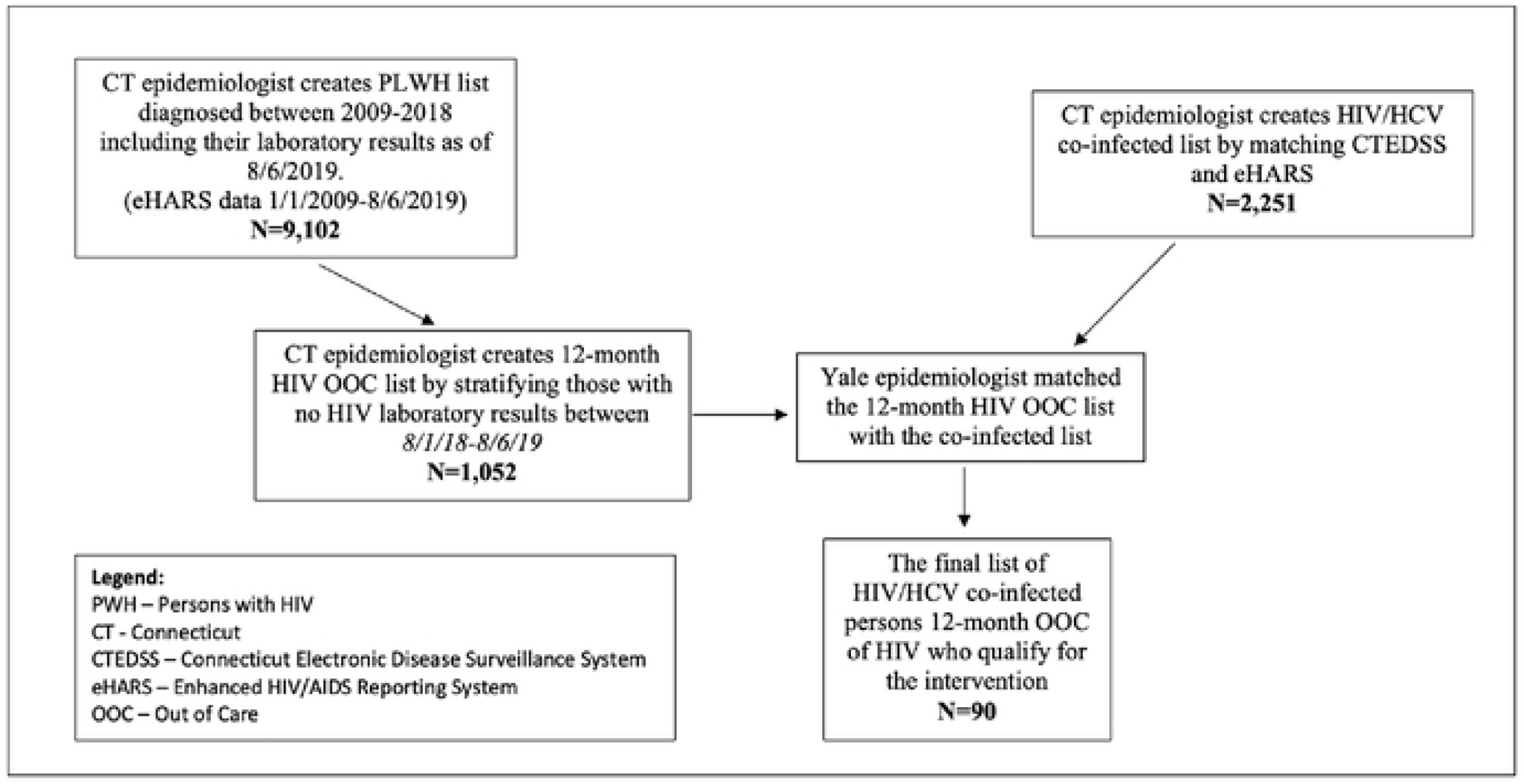
Surveillance data flow for identification of Persons with HIV (PWH) who qualify for the Data to Care intervention

Using the same eHARS list, the CT DPH epidemiologist created a list of persons with HIV (PWH) who were diagnosed with HIV prior to or including 2018 which included their most recent HIV laboratory results (eHARS data from 1/1/2009 to 8/6/2019). Persons who received an HIV diagnosis in 2019 were excluded since a full year is needed to finalize and confirm HIV data. The CT DPH epidemiologist created the 12-month PWH OOC list to include persons who had no HIV laboratory results from 8/1/2018 to 8/6/2019. The Yale epidemiologist generated the 12-month HIV/HCV co-infected OOC cohort list by matching the PWH OOC list to the HIV/HCV co-infected list. Before finalizing the list, HCV data were evaluated using CTEDSS to exclude anyone with evidence of HCV viral clearance. (as defined by positive HCV PCR followed by negative PCR).

### Process for Identifying, Locating, and Re-engaging HIV OOC Persons with HIV/HCV Co-infection

#### List generation

After generating the list of eligible PWH as noted above, a tracking log (an Excel spreadsheet consisting of patient identifying information, HCV and HIV laboratory details) was created for recording outcome data, that was manually filled in by the DIS supervisor and DIS staff. Using this list, three phases of further data cleaning and assignment to DIS workers began (see **Figure 2**). Efforts began 9/1/2019 and continued until 3/31/2020.

**Figure 2:**
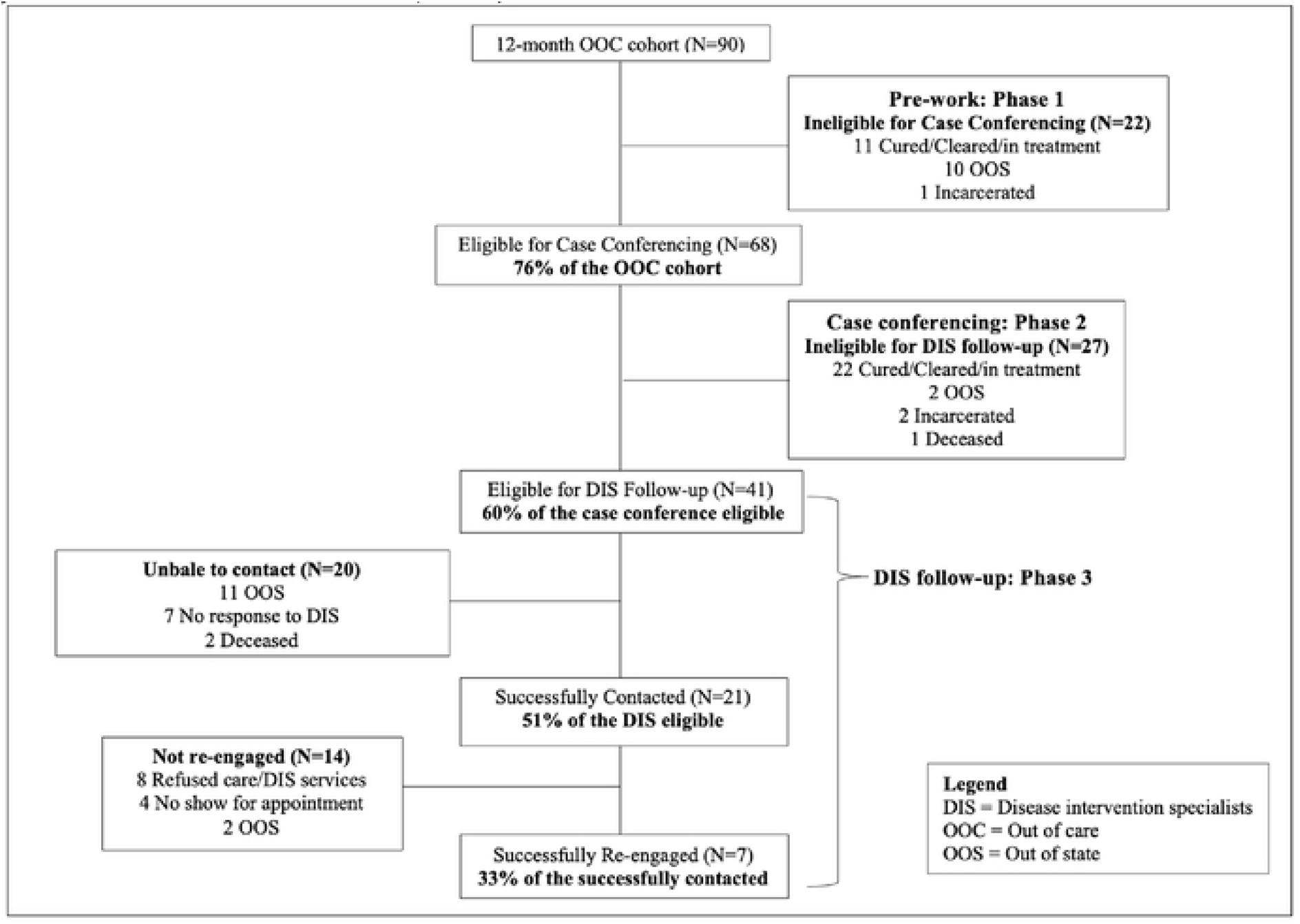
Consort diagram showing eligibility for the phases of the Data to Care intervention for the 12-month OOC Cohort (N=90)

#### Phase 1-Pre-Work

The goal of Phase 1 was to use available DPH data systems to streamline the co-infected lists to include persons who were alive, residing in CT, not treated for HCV. The DIS supervisor used the Department of Corrections (DOC) database, LexisNexis, and CTEDSS (data updated regularly throughout the project) as ancillary databases to assess eligibility for Phase 2 case conferencing. Persons who were deceased, incarcerated, out of state (OOS) residents, and those with at least one negative PCR result reported through CTEDSS (indicating prior successful treatment or spontaneous clearance) were not eligible for Phase 2. Persons lacking a provider or sufficient contact information were eligible for Phase 2.

#### Phase 2-Case Conferencing

The goal of Phase 2 was to solicit clinic-level information through a direct conversation between the DPH and clinic staff (case conferencing). The DIS supervisor spoke with relevant clinic staff who reviewed medical records to verify if their patients were truly OOC for HIV and to ascertain their HCV treatment status since data in CTEDSS were not always up-to-date due to data backlogs. If needed, the DIS supervisor contacted eligible patients’ last known HIV provider obtained from eHARS by phone. The ensuing discussions yielded updated outcomes that were recorded in the tracking log. Persons eligible for active DIS follow-up (Phase 3) included patients lost to follow-up by the clinic and those with insufficient information to determine eligibility.

#### Phase 3-DIS Follow-Up

The goal of Phase 3 was active case-finding by DIS workers. The CT DPH employed three dedicated DIS to locate and contact patients needing care engagement including those OOC for HIV. DIS tracked assigned patients; assessed barriers to care after contact, provided HIV and HCV brief treatment education, and facilitated re-engagement with the HIV clinic. Patients were actively followed-up for 30 days once DIS efforts began. If a patient was contacted, the DIS would work with the patient until they were re-engaged (defined as making a provider appointment). If after 30 days there was no response to DIS, passive efforts (awaiting responses to initial contact efforts) would continue for another 60 days.

Outreach methods followed a dedicated protocol (see Appendix) and included phone calls, text messages, emails, field visits, and letters with most recent contact information supplemented by clinic records; use of these methods was not mutually exclusive, e.g. if needed, phone calls and field visits were used with the same patient. Tabulation of outreach efforts was designated by final outcome, e.g. persons who were successfully re-engaged had relevant outreach efforts attributed to that outcome. Progress and outcomes for Phase 3 were recorded in REDCap, a secure web-based electronic data capture tool hosted at Yale University.

#### Outcome Definitions

1. Successful contact – DIS reached a patient to schedule a clinic appointment.
2. Successful re-engagement – patient had either a scheduled appointment or clinic visit with an HCV treatment provider by the end of study. The re-engagement success rate is calculated as the ratio of those re-engaged to those successfully contacted.
3. Cure-clinic or surveillance based evidence of SVR (sustained virologic response defined as HCV PCR negative 12 weeks post treatment).

### Project Variables

Surveillance variables included first name, last name, date of birth, birth sex, race/ethnicity, HIV transmission category and most recent available HIV viral load (VL) results and date.

### Statistical Analysis

Descriptive analyses were performed to examine the distribution of demographic and HIV clinical information variables for each OOC cohort observed during each project phase. The variables included birth year, age, birth sex, race/ethnicity, HIV VL level, and HIV transmission category. Descriptive analyses were also performed to examine the outreach methods used and time spent by DIS for patient follow-up.

### Ethical Considerations

This project was approved by the Yale IRB (protocol #2000025960) and the CT DPH Human Investigations Committee (protocol #914). The CT DPH Human Investigations Committee gave permission for a Yale researcher (MW) to only view identifiable information while physically at the DPH and no data was accessible outside the DPH. All study data were located and secured at the CT DPH according to their internal policies.

## RESULTS

### Demographics

As shown in **Table 1**, there were 90 HIV/HCV co-infected patients who were OOC for HIV (66% male, 60% born prior to 1965, 44% Hispanic, 71% persons who inject drugs (PWID), 74% with most recent HIV VL level undetectable). Out of these, 41 were eligible for DIS follow-up. Compared to the total OOC, those eligible for DIS follow-up had greater percentages of persons born in 1965 or after (44% vs 40%), Hispanic (54% vs 42%), and detectable HIV VLs (34% vs 26%).

**Table 1:** Demographics and clinical characteristics of persons with HIV/HCV co-infection stratified by HIV Out of Care and eligibility for Disease Intervention Specialist Services.

| Variable<sup>†</sup> | Category | Out of Care<sup>¶</sup><br>(N=90) |  | DIS<br>Eligible<sup>††</sup><br>(N=41) |  | Successfully<br>contacted<sup>‡‡</sup><br>(N=21) |  | Successfully<br>re-engaged<sup>§§</sup><br>(N=7) |  |
| --- | --- | --- | --- | --- | --- | --- | --- | --- | --- |
|  |  | N | % | N | % | N | % | N | % |
| Sex | Male | 59 | 66% | 27 | 66% | 16 | 76% | 3 | 43% |
|  | Female | 31 | 34% | 14 | 34% | 5 | 24% | 4 | 57% |
| Birth Year | Prior to 1965 | 54 | 60% | 23 | 56% | 10 | 48% | 3 | 43% |
|  | 1965 and after | 36 | 40% | 18 | 44% | 11 | 52% | 4 | 57% |
| Race/Ethnicity<sup>‡</sup> | White | 25 | 28% | 9 | 23% | 4 | 20% | 1 | 14% |
|  | Black | 25 | 28% | 9 | 23% | 7 | 35% | 4 | 57% |
|  | Hispanic | 38 | 44% | 22 | 54% | 9 | 45% | 2 | 29% |
| HIV Transmission<br>Category | PWID | 64 | 71% | 28 | 68% | 14 | 67% | 5 | 71% |
|  | Other<sup>§</sup> | 26 | 29% | 13 | 32% | 7 | 33% | 2 | 29% |
| HIV Viral Load<br>Level | Detectable ($\geq 200$) | 23 | 26% | 14 | 34% | 8 | 38% | 2 | 29% |
| | Undetectable ($< 200$) | 67 | 74% | 27 | 66% | 13 | 62% | 5 | 71% |

Note: DIS = disease intervention specialists; PWID = persons who inject drugs
<sup>†</sup> Calculated with unknowns excluded
<sup>‡</sup> Does not include other or multiple race categories due to small sample sizes
<sup>§</sup> Includes: heterosexual, Men who have sex with men (MSM), MSM with PWID, and Other transmission categories
<sup>¶</sup> Includes persons with no HIV laboratory tests between 8/1/2018-8/6/2019 and matched to DPH HIV/HCV co-infected list (based on matching eHARS (from 1/1/2009 to 8/6/2019) and CTEDSS (from 1/1/1994 to 8/17/2019))
<sup>††</sup> Persons eligible for active DIS follow-up included patients lost to follow-up by the clinic and those with insufficient information to determine eligibility.
<sup>‡‡</sup> DIS reached a patient to schedule a clinic appointment.
<sup>§§</sup> Patient with a scheduled appointment (excluding no shows) by the end of study or a completed visit with an HCV treatment provider

### Role of Pre-Work and Case Conferencing Phases to Determine DIS Eligibility

**Figure 2** llustrates the data cleaning role of the pre-work (Phase 1) and case-conferencing (Phase 2). Of the 90 OOC persons, 68 (76%) were eligible for case conferencing; 22 persons (24%) were deemed ineligible for case conferencing based on data clean-up from surveillance and other DPH data. Among those undergoing case conference (N=68), 27 (39.7%) were ineligible for DIS follow-up. Patients who were already cured/cleared/in treatment were the most common reason for case confercning (11, 50%) and DIS follow-up (22, 81%) ineligibility.

### DIS Re-engagement Outcomes

Figure 2 shows that among those eligible for DIS (N=41), 20 (48.8%) were not successfully contacted (11 patients residing out of state (55%); 7 (35%) failing to respond to DIS attempts at communication; 2 (10%) were deceased). Among the successfully contacted (N=21), 7 (33%) were successfully re-engaged; 14 (66.7%) were not re-engaged of which 8 (38%) refused DIS services/clinical care, 4 (19%) were appointment no-shows, 2 (9.5%) were out of state. Overall, no patients achieved SVR by study’s end.

### Analysis of Successfully Contacted and Re-Engaged

**Table 1** compares the DIS-eligible group to persons who were successfully contacted and re-engaged. While males accounted for the majority of DIS-eligible and successfully contacted, females represented the majority (57%) of those successfully re-engaged. While the majority (56%) of DIS eligible were born prior to 1965, the majority of successfully contacted and re-engaged were born after 1965. PWID constituted the majority of DIS-eligible (68%) as well as persons successfully contacted and re-engaged. While Hispanic persons were the majority group (54%) in the DIS-eligible, Black persons comprised the majority (57%) of those successfully re-engaged. The majority of DIS-eligible persons as well as successfully contacted and re-engaged had undetectable HIV VLs (at last measure prior to being OOC).

### DIS Outreach Modalities

**Table 2** shows DIS outreach tools and the estimated time DIS spent (based on approximate average time reported) with the 41 eligible patients. Of 192 total outreach attempts, phone calls were used most frequently (59% of the total attempts); field visits accounted for the greatest follow-up time (77% of total minutes spent). Other methods include the combination of mailed letters, text messages, and social media messages which contributed 12% of tools used and 3% of follow-up time spent. These effort distributions were also reflected among those successfully contacted and re-engaged where phone calls were used most often but field visits took the most time. Active follow-up on those ultimately re-engaged (N=7) took 870 minutes or 124.3 minutes per patient. For the not re-engaged, the total time was 2,885 minutes cumulatively or 206 minutes per patient. Compared to the re-engaged, DIS spent more time conducting field visits for the not re-engaged (78% vs 64% of total minutes). For those who were re-engaged, DIS spent more time with phone calls (29% vs 19%) compared to those not re-engaged.

**Table 2:** Distribution of outreach attempts and time spent stratified by outreach tools used for disease intervention specialist (DIS) follow-up.

| DIS tools | DIS eligible (N=41)<sup>†</sup> |  | Successfully contacted (N=21)<sup>‡</sup> |  | Re-engaged (N=7)<sup>§</sup> |  | Not re-engaged (N=14)<sup>¶</sup> |  |
| --- | --- | --- | --- | --- | --- | --- | --- | --- |
|  | Total attempts (N=192) | Total Minutes (5,807) | Total attempts (N=137) | Total Minutes (3,755) | Total attempts (N=39) | Total Minutes (870) | Total attempts (N=94) | Total Minutes (2,885) |
| Phone calls (Avg. 10 mins) | 114 (59%) | 1,140 (20%) | 81 (59%) | 810 (22%) | 25 (64%) | 250 (29%) | 56 (60%) | 560 (19%) |
| Field Visits (Avg. 80 mins) | 56 (29%) | 4,480 (77%) | 35 (26%) | 2,800 (75%) | 7 (18%) | 560 (64%) | 28 (33%) | 2,240 (78%) |
| Other<sup>††</sup> (Avg. 8.5 mins) | 22 (12%) | 187 (3%) | 21 (15%) | 145 (4%) | 7 (18%) | 60 (7%) | 10 (7%) | 85 (3%) |
| <sup>†</sup> Persons eligible for active DIS follow-up included patients lost to follow-up by the clinic and those with insufficient information to determine eligibility<br><sup>‡</sup> DIS reached a patient to schedule a clinic appointment<br><sup>§</sup> Patient with a scheduled appointment (excluding no shows) by the end of study or a completed visit with an HCV treatment provider<br><sup>¶</sup> Patient with no scheduled appointment by the end of study<br><sup>††</sup> Other includes text messages, mailed letters, and social media messages |  |  |  |  |  |  |  |  |

## DISCUSSION

We piloted a D2C strategy leveraging HIV OOC status to identify and re-engage persons also infected with HCV who lacked HCV treatment. Using surveillance data, clinic case conferencing, and DIS field work, we found that efforts resulted in intensive data clean-up including updating HCV cure status and re-engagement follow-up activities but failed to achieve additional HCV treatment initiations and cures.

This study highlights challenges associated with outreach within a population defined as OOC for HIV, namely lacking HIV surveillance laboratory testing in the preceding 12-months. Given that available, timely data is confounded by inherent surveillance data lags (generally one year lag for data finalization), the OOC duration in this definition may actually be 24-months or more.

Our methodology for D2C highlights the important role of this approach for data cleaning. In the pre-work and case conferencing phases, we found that 33 patients (37% of the 12-month OOC cohort) were HCV cleared/cured/in treatment, showing that being HIV OOC did not necessarily preclude treatment for HCV. This clean-up process illustrates that the current HCV surveillance database is incomplete, with gaps in ascertainment of cures that are likely due to temporal lags in populating the database and gaps in CT DPH’s Electronic Lab Reporting (ELR) interface.

Another challenge was the lack of updated and accurate patient status and contact information within public health surveillance databases. Investigation efforts at each phase led to discoveries of redundant reasons for DIS ineligibility. For example, patients that DIS could not contact included deceased or out-of-state residents; ideally this should have been captured during the pre-work phase. Many were DIS ineligible due to lack of reliable contact or provider information. These determinations were critical in discerning which patients were actually eligible for DIS re-engagement strategies. Overall, most re-engagement activities were spent excluding 77% of the initial OOC population. This finding has been highlighted in other D2C efforts, revealing the extent of data inaccuracy, possibly overestimating the number of OOC persons.^26–29^ In fact, surveillance data clean-up is an important function of D2C, especially when defining who is OOC. In another D2C study, we found that case conferencing with HIV clinics revealed PWH identified to be recently OOC by surveillance data (no HIV laboratory results for 6 months) were actually in care (71%) as defined by the clinics. This refinement of care status would enable the DIS to more efficiently focus their resources on an at-risk group needing intensive follow-up.^30^

For those ultimately deemed DIS-eligible, the workload of the DIS was considerable. Most of the time was spent on field visits (77%) followed by phone calls (20%). We found that once contact was established, the DIS spent more time following-up with non-re-engaged patients compared to re-engaged (206 vs 124 minutes per patient), concluding this population is not only hard to treat but hard to persuade. The low re-engagement yield in this study may have been due to barriers faced by this long-term OOC population that were simply not amenable to the short turnaround nature of DIS activities. In other work, we found that successful facilitators of promoting HCV care are often based on long-term trusting relationships with clinic providers and go beyond single re-engagement at a clinic visit.^29^ We would predict this D2C strategy of using DIS to locate and attempt to re-engage OOC HIV/HCV co-infected persons was neither time nor cost efficient. We also caution again using field visits as they took the greatest amount of time and were no more effective than the other tools for this patient population.

Previous studies focusing on engaging hard to treat populations into HCV care have looked at various barriers and facilitators.^30–34^ Barriers within the clinical realm have undergone re-evaluation. For example, extensive pre-treatment evaluation and treatment monitoring have been addressed with simplified treatment approaches.^14–16, 18, 20^ Telehealth interventions may be ideal for addressing some of these issues.^35^ These approaches, while important, may not be adequate for difficult to reach populations which constitute the HCV cure gap, highlighting personal, interpersonal, institutional, and social barriers.^31^ Because our numbers are low, we were unable to identify clear demographic predictors for persons who can be successfully re-engaged; PWID, Black, female, born after 1965 were among this small group. Interestingly, while the majority of persons successfully re-engaged had undetectable HIV VLs at last measure, it is unknown whether they remain virally suppressed as this group has been OOC for 12 months. Our previous studies looking at the HCV viral clearance cascade on the statewide level and within HIV clinics suggest that a lack of recent HIV viral suppression is a marker for lack of engagement, correlating with lack of HCV care engagement.^19, 36, 37^ Other factors such as treatment literacy, media representations, trusted and steady provider relationships, perceived lack of adherence support, concerns for treatment cost, were potential self-identified barriers to engagement in another study.^38^ Often, patients do not enter HCV treatment due to multiple concurrent barriers,^39^ suggesting that the D2C approach using DIS workers which is inherently short-lived and focused on brief educational efforts and motivational interviewing, may not be sufficient to overcome such longstanding and complex issues.^31^ Additionally, active substance use issues may need to be addressed, which is beyond the scope of DIS.

Finally, this study presents an interesting perspective on how the traditional HCV treatment models may not best serve all patients, especially those at greater risk of becoming OOC. Decentralized and simplified HCV treatment approaches including low-barrier rapid start programs which capitalize on the availability of well-tolerated pan-genotypic regimens, have been successfully piloted, e.g. in young PWID.^40^ We speculate that D2C, a public health approach that is inherently constrained by its retrospective data-focus, can potentially be combined with rapid start programs to “jump start” HCV treatment in challenging populations such as those encountered in this study.^40–41^ Finally, innovative approaches beyond telephone calls, texts and field visits which use community networks may provide alternative effective re-engagement strategies.

### Limitations

Study numbers were low, so it was not possible to make correlations with outcomes. This study was conducted during the early stages of COVID-19 pandemic which may have limited the success rate of re-engagement efforts. We did not account for time spent in Phase 1 and Phase 2 which was expended primarily by the DIS supervisor, prior to assignment to DIS for case finding; this would have given additional information on time expenditures for this intervention.

### Strengths

This study is one of the first to examine D2C re-engagement efforts for PWH co-infected with HCV in need of HCV treatment. It leverages previous D2C DIS infrastructure designed for and funded by HIV surveillance efforts. This approach promoted HCV treatment education by DIS in the field. Surveillance-based data extraction to identify OOC cohorts allowed state-wide evaluation, proving more efficient than individual clinics creating and validating their own OOC lists. Surveillance data use allows for reproducibility by other health departments and the case conferencing methodology allowed the DPH to foster partnerships with clinics, leading to future public health improvements.^30^

## CONCLUSIONS

We successfully leveraged HIV D2C processes to identify and characterize HIV/HCV co-infected persons who were long term OOC for HIV who appeared to lack HCV treatment. We used a 3 phase approach based on DPH and clinic input that enabled surveillance data clean-up and identification of persons who were eligible for case finding by DIS. The intensive workload for DPH staff including DIS resulted in a moderate degree of successful re-engagement but no additional cases of HCV cure. To compliment D2C approaches, future studies should evaluate D2C cost-effectiveness and enable incorporation of clinic- and community-based interventions that address ongoing and individualized patient barriers and promote rapid, low-barrier treatment approaches.

## Data Availability

The data underlying the results presented in the study are available from the Connecticut Department of Public Health (CT DPH). Please contact (860) 509-7900.

## Acknowledgements

Special thanks to Venesha Heron, Reina Cordero, and Emily Hartwell from the CT DPH for their Disease Intervention Specialists (DIS) work and Rodolfo Lopez for his assistance. We also thank the clinic staff from participating HIV clinics for their help with case conferencing.

The Department of Public Health Human Investigations Committee approved this research project, which used data obtained from the Department of Public Health. The Department of Public Health does not endorse or assume any responsibility for any analyses, interpretations or conclusions based on the data. The author assumes full responsibility for all such analyses, interpretations, and conclusions.

